# Diurnal Temperature Variation and Using Fever to Screen for Infectious Diseases

**DOI:** 10.1101/2020.08.04.20164095

**Authors:** Aaron C. Miller, Scott Koeneman, Philip M. Polgreen

## Abstract

Fevers have been used as marker of disease state for hundreds of years and are frequently used to screen for infectious diseases during infectious disease outbreaks. However, body temperature and fevers have been shown to vary over the course of a day and across individuals by age, sex and other characteristics. The objective of this paper is to describe the individual variation in diurnal temperature patterns during episodes of febrile activity using a database of millions of recorded temperatures across the United States. We then model the probability of recording a fever during a single reading at given time for individuals who are experiencing a febrile episode. We find a wide variation in body temperatures over the course of a day and across individual characteristics. Similarly, the likelihood of recording a fever may vary widely by the time of day when the reading is taken and by an individual’s age or sex. These results suggest diurnal temperature variation and demographics should be considered when using body temperature to screen for disease, especially for diseases that are contagious.

## Background

Fever has been a hallmark of disease for thousands of years. The relationship between fevers and the disease state was first described in Akkadian cuneiform inscriptions thousands of years prior to the first thermometer.^1,2^ The ancient Greeks described the correlation between temperature and pulse^3^, and given the ability to perform tactile measurements of warmth, the measurement of pulse was often used as a proxy for a fever. Galileo invented the thermoscope in the 1590s, and soon after, this device was used to determine normal and abnormal temperatures in humans^4^. In 1600s Santorio Santorio coined the term “thermometer”, and was also one of the first to appreciate the clinical value of temperature records.^5^ The invention of the mercury thermometer by Fahrenheit in 1714, allowed for more widespread measurements of fevers. Substantial thermometer measurements of humans were recorded by DeHaen in 1773, who noted diurnal fluctuations, and differences in readings among people of different ages and with different diseases.^4^ Problems with variation and calibration of thermometers inhibited their widespread use for medical applications for almost a hundred years. However, in 1851, Wunderlich started his landmark work in thermometry taking and recording multiple temperature measurements for thousands of different patients in the hospital where he worked.^6^

In Wunderlich’s comprehensive examination of thermometry, He established that temperatures change across the spectrum of ages.^6^ He is credited with establishing the normal temperature of a healthy adult as 37°C, or 98.6°F and in the process of his descriptive work, he established a clear diurnal pattern of temperatures and demonstrated that normal temperatures represent a range rather than an actual value. He established the notion of an objective fever measurement. He also showed that temperatures varied between mem and women and that older adults had lower temperatures than children.^7^

While Wunderlich was not the first to describe the diurnal pattern of temperatures,^4^ his authoritative work demonstrated a clear pattern with lower temperatures in the morning that increase in the early evening, a pattern that has been replicated multiple times since. This diurnal pattern exists for healthy individuals as well as for subjects suffering from a wide range of infections. For example, a majority of patients with malaria,^8,9^ and other infections including tuberculosis, pneumonia, endocarditis and urinary tract infections^10^ have temperatures that peak in the evening.

Specific patterns of fever, with a few exceptions, are not generally helpful for differentiating different infections from each other.^10^ Furthermore because fevers are part of an inflammatory response, they occur with non-infectious conditions including malignancies and autoimmune diseases and are also subject to diurnal patterns.^10^ However, the fact that diurnal patterns exist as part of an inflammatory response has great clinical and public health importance. Because of this diurnal temperature pattern, taking the temperature at the wrong time of day may decrease the likelihood of detecting a fever. Given that the detection of fevers is important for both clinical and public health purposes (e.g., screening of workers or travelers during an outbreak), it is important to understand how the diurnal temperature pattern effects the ability of screening programs to detect fevers.

The purpose of this paper is to describe the diurnal variation in temperatures patterns using a large database of millions of recorded temperatures, and to build a model of temperature to determine the probability of detecting a fever during a febrile episode across different age groups, depending on the time of day.

## Methods

### Data

We use temperature readings recorded from Kinsa Smart Thermometers. These thermometers connect to, and pair readings with, a smartphone application. The thermometer is commercially available across numerous retail locations and thermometers have also been made available to schools, parents and families with children through the Kinsa school program.^11^ There are hundreds of thousands of thermometers located across the United States. We use temperatures recorded from 01/27/2014 through 07/15/2019; these data have previously been shown to be highly correlated with influenza-like-illness (ILI) activity, and temperature readings have been used to generate national, state and local forecasts of illness activity.^12,13^

### Constructing Illness Episodes

We construct *illness episodes* by first grouping temperature readings into clusters of activity that are likely to represent a period of illness in a particular device user. In prior work, we demonstrated this approach for collapsing temperature readings recorded over a series of days, and these episodes were used to identify illness duration, household transmission events, and biphasic fever patterns (two febrile-illness episodes that occurred in short succession).^12^ Each of these illness patterns were also shown to be highly correlated with ILI activity.

All readings recorded by the same user profile on a given device, that occur within a fixed interval of another reading are grouped into an episode. In this analysis, we use 24 hours as the maximum time between readings to define an episode. Because some users may take recurrent readings on a regular basis (e.g., fertility planning), we exclude episodes that last longer than a typical illness (i.e., 7 days). Finally, we define illness episodes to be those episodes where at least one fever (temperature ≥ 100.4 F°) was recorded during the episode interval. The following analysis describes diurnal temperature patterns among readings in these identified illness episodes. We also conduct a sensitivity analysis where we consider 12, 36, 48 and 72 hours as the maximum time between readings in a given cluster.

### Constructing a representative healthcare workforce

We analyze the likelihood of recording a fever during an illness episode at a given point in the day for a typical healthcare worker. To construct a representative healthcare workforce, we used the distribution of ages of workers employed in the healthcare sector from the Bureau of Labor Statistics (see: https://www.bls.gov/cps/cpsaat11b.pdf). The following table, **Appendix Table 1**, provides the proportion of healthcare workers in each corresponding age group. We scale fever readings by these proportions to generate a pattern of temperature taking representative of a typical healthcare worker, on average.

### Statistical Analysis

To estimate mean temperature, we used a least-squares regression model with a sinusoidal time component, and interaction terms for age group and sex. Four sinusoids were used: sine and cosine terms with 12-hour periodicity, and sine and cosine terms with 24-hour periodicity. This was done to match the oscillating pattern observed in the descriptive plots, while enforcing periodicity. Each of these four terms was interacted with age group and sex, which were also interacted with each other. Note: that the baseline case for the model is a female in age group 0-15, which is reflected in the model coefficients.

To estimate the probability of recording a fever during a given time period, we use a generalized linear model with a log-link and binomial distribution, along with the same sinusoidal component as the mean temperature model, interacted with age and sex.

Finally, we estimate both the mean temperature curve and the probability of recording a fever for a representative healthcare population. We weight parameter estimates using the breakdown of age-specific employment percentages from the BLS, and use the delta method to estimate the confidence interval around these estimates.

#### Sensitivity analysis

We conduct a sensitivity analysis by exploring multiple ways to define an illness episode. Because illness episodes were defined by the time between readings and a registered fever, it may be the case that some of the non-febrile readings we include occurred just outside the illness period. Thus, we use the approach described above (based on a 24-hour window) to model mean temperature while using 12, 36, 48 or 72 hours also alternative timeframes to define a period between recurrent readings that are included in a single episode of readings. We compare these models’ predictions to those obtained from the 24-hour period definition.

## Results

There were 10,263,424 total temperature readings across the study period. After grouping readings into periods of multiple-day temperature taking, we identified a total of 1,856,988 episodes. Of these, a total of 540,870 episodes involving at least one febrile reading, and these episodes were comprised of a total of 3,653,518 temperatures. Each fever episode had a mean and median of 6.75 and 4 readings, respectively.

### The diurnal temperature pattern

There is a natural diurnal pattern in body temperatures that peaks in the evening (around 7-9 pm) and is lowest in the early morning (between 6-8 am). This pattern has been previously described in multiple investigations, and can be observed simply by computing the mean temperature during any given minute of the day. **Figure 1** depicts this diurnal pattern by age group. Here each point represents the mean temperature of all readings recorded during a given hour of the day. The blue curve depicts the smoothed trend, using a loess curve. This diurnal pattern can be seen to vary by age and time of recording. In general, the mean temperature in the diurnal curve decreases with older ages. **Appendix Figure 1** depicts the same trend as **Figure 1**, but in **Appendix Figure 1** we have broken out the mean of all readings by each minute of the day. The distribution of temperatures recorded at any given time point (minute or hour) is approximately Gaussian. **Appendix Figure 2** depicts histograms of temperature recorded for each hour of the day and **Appendix Figure 3** depicts a kernel density of temperatures recorded each hour of the day.

**Figure 1:**
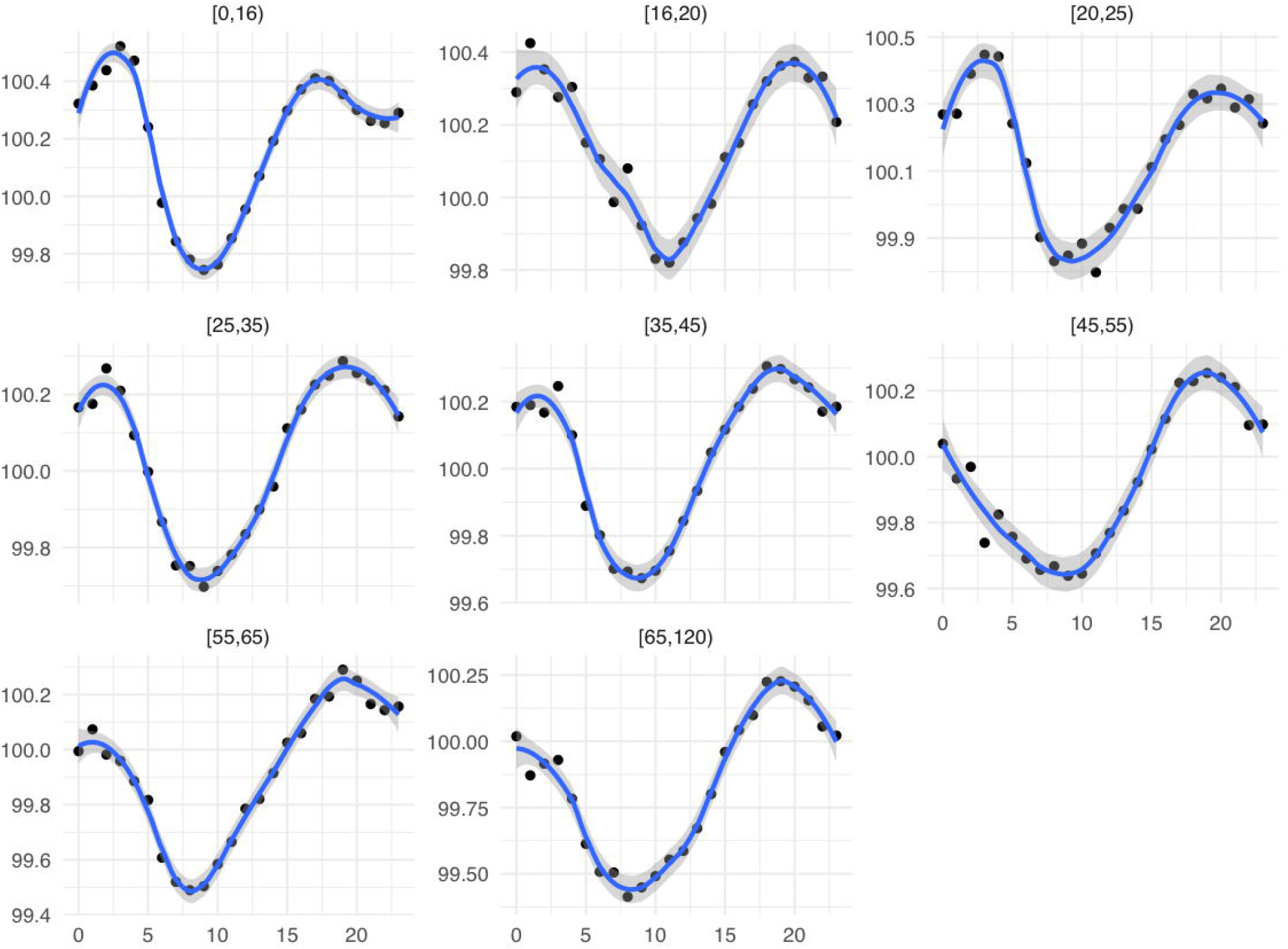
Mean temperature by hour of the day aggregated across all readings in a given age group. Each individual plot depicts a particular age group. The blue line gives the smoothed trend.

**Figure 2:**
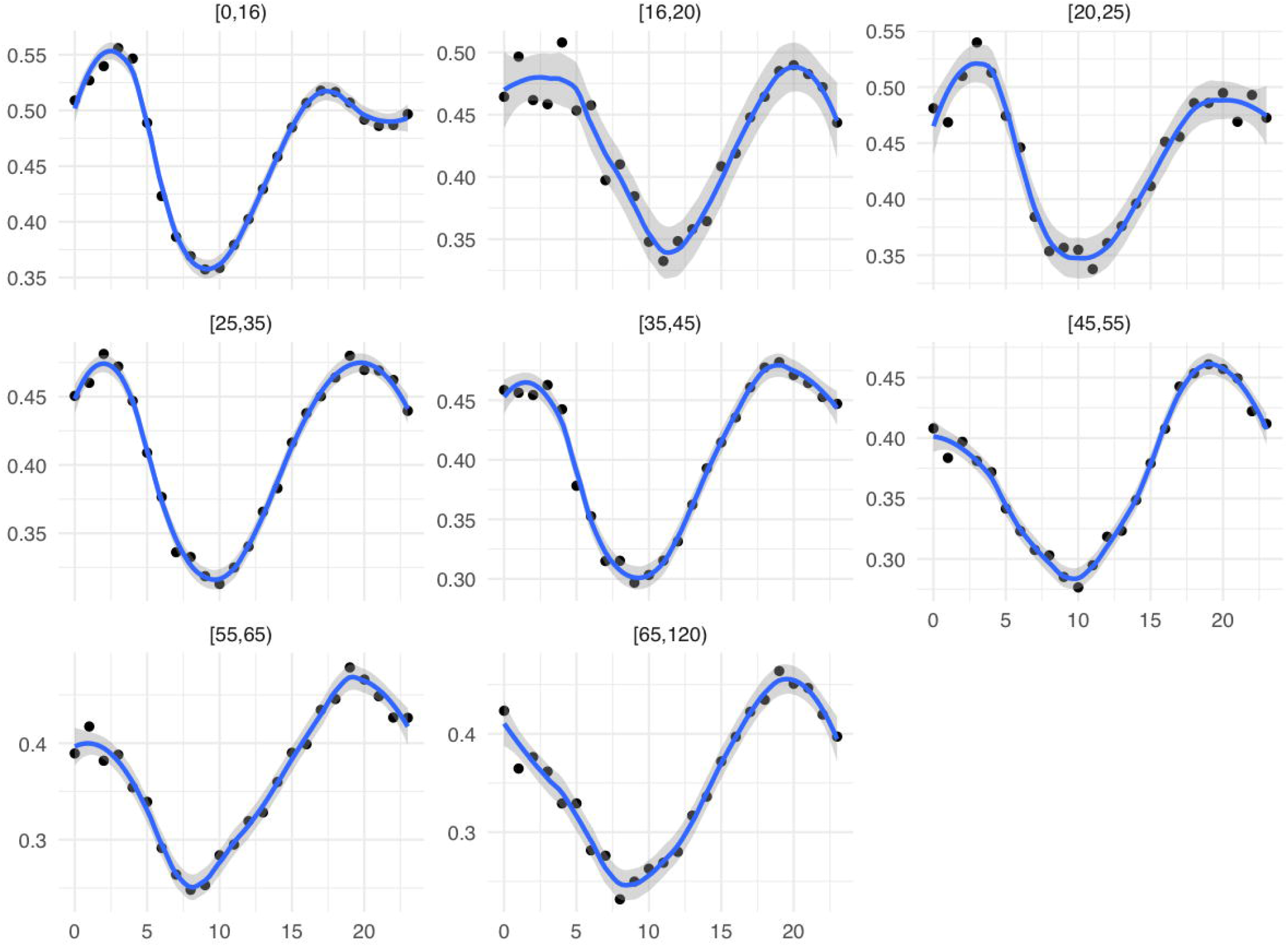
Probability of recording a fever during an illness episode for each hour of the day, for different age groups. Each plot depicts the trend for a particular age group.

**Figure 3:**
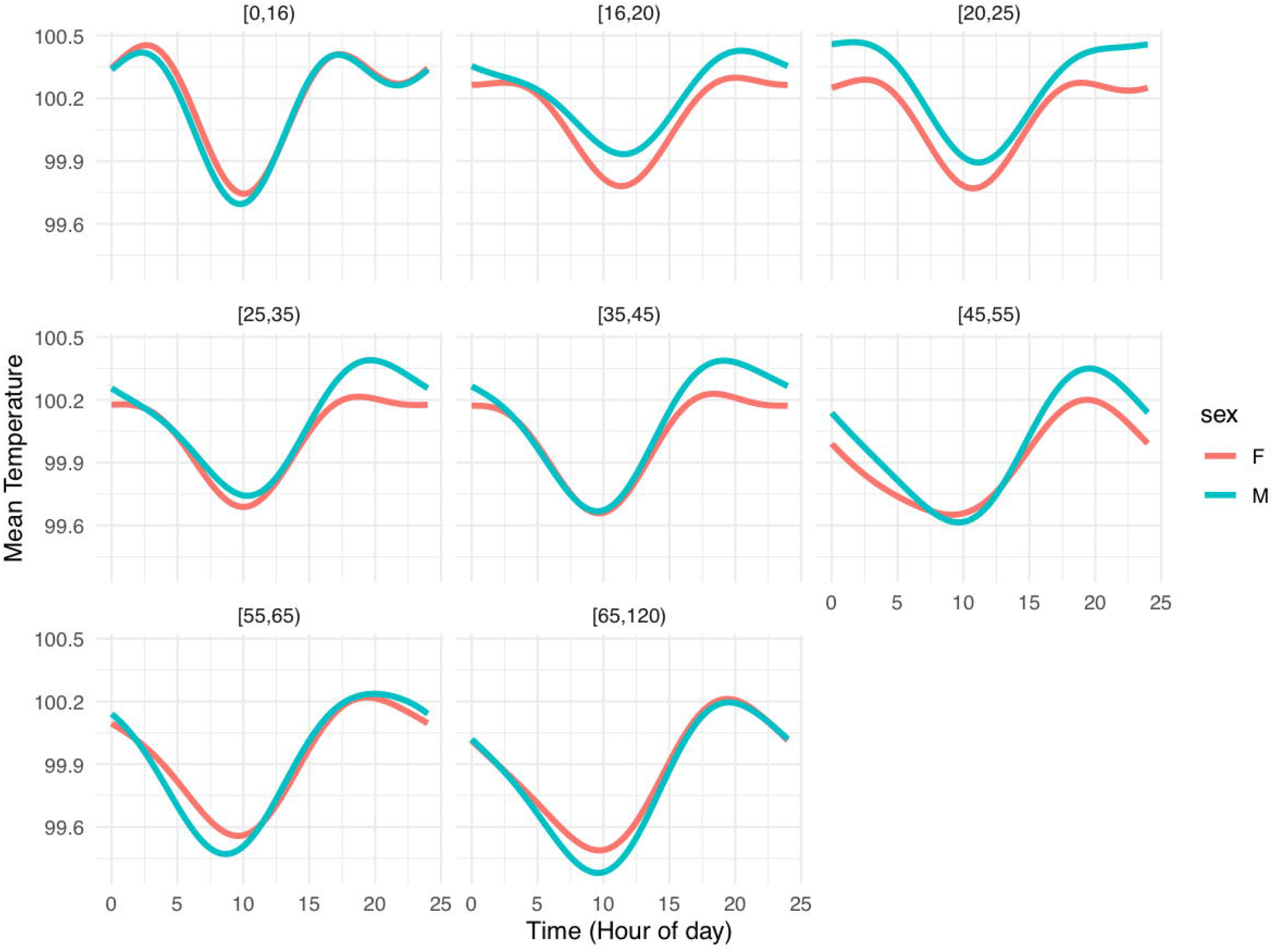
Diurnal temperature pattern for mean temperature by time of day using the sinusoidal regression model. Trends are depicted by age group and broken down by sex.

We can ask the question *how likely are we to draw a fever reading (>100F) at any given hour during the day*. Using the temperatures recorded for each age group, we compute the probability of recording a temperature >100.4F. We simply calculate the proportion of all temperature readings that were febrile during a given hour of the day for a specific age group. **Figure 2** depicts this probability by hour for each of the age groups.

### Modeling the diurnal pattern

We use the sinusoidal model described above to estimate the mean diurnal pattern by time of day, while controlling for age and sex. The diurnal pattern for different age groups and by sex is depicted in **Figure 3**; the remaining model coefficients are given in **Appendix Table 2**. We see that, similar to the univariate descriptive plots for all demographics, temperature during an illness episode tends follow a cyclical pattern. Mean temperatures decrease until a certain point in the morning, hitting a trough, and then increasing until a certain point in the evening, hitting a peak. The overall curve tends to be lower for those in more advanced age groups, while for males age 16-55, mean temperature throughout the day tends to be higher than that of females. This does not appear to be the case for individuals younger than 16 or older than 55.

### Modelling probability of observing fever by time

We use a similar approach as above to estimate the probability of observing a fever during a given time of the day. To estimate the probability of observing a fever we use a generalized linear model with a logit link and binomial distribution. In **Figure 4**, we have plotted fitted values for each time period by age group and sex using the sinusoidal time component described above. Consistent with the mean temperature model, the diurnal pattern varies in a cyclical form with the time of day. Depending on the particular age and sex of an individual, the probability of recording a fever during an illness episode may vary considerably by the time of the day. For example, a male individual over the age of 65 years has around a 30% chance of registering a fever at around 9:00 am but a 55% chance of registering a fever at 8:00 pm.

**Figure 4:**
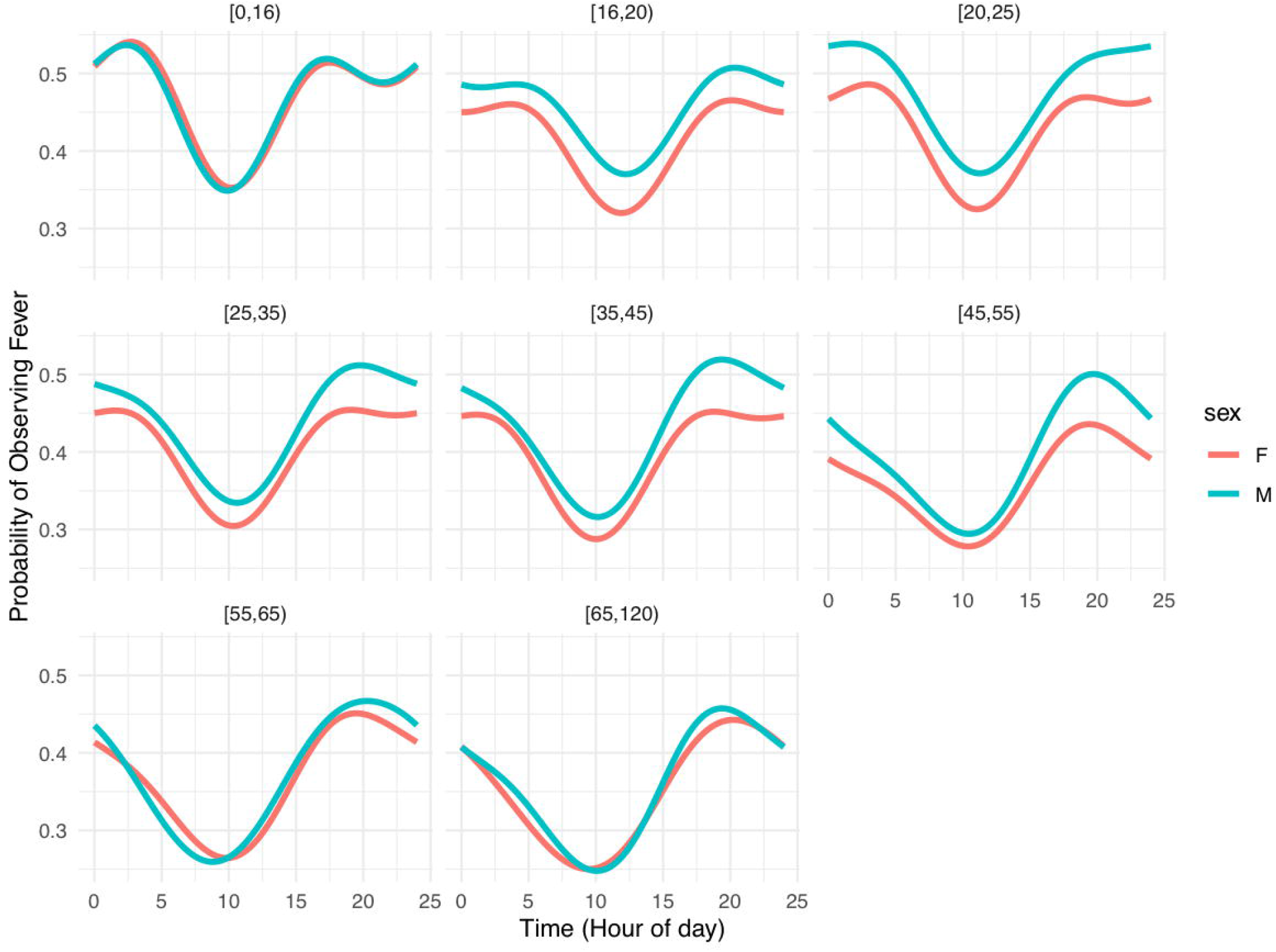
Diurnal temperature pattern for probability of recording a fever as a function of the time of day using the sinusoidal regression model. Trends are depicted by age group and broken down by sex.

Model coefficients for the fever-probability model are given below, in **Appendix Table 3**. We see that the probability predictions follow the same trends observed in the mean temperature model, with a trough in the morning and a peak in the evening. Those in advanced age groups also tend to have lower probability of a fever in general, while males ages 16-55 are slightly more likely to have a fever than females.

### Temperature in a healthcare workforce

Using the estimates from the fever-probability model, in Table 3, we can construct the probability of drawing a fever during any given hour of the day for a representative healthcare worker population. We weight the coefficient estimates in **Appendix Table 3** by the proportion of each age group in the overall healthcare workforce, described in Table 1. **Figure 5** depicts these weighted probability values for any given hour during the day. The probability of recording a fever from an illness during a given time of the day for a typical healthcare worker, ranges from around 36% at 9 am to around 54% at 7 pm.

**Figure 5:**
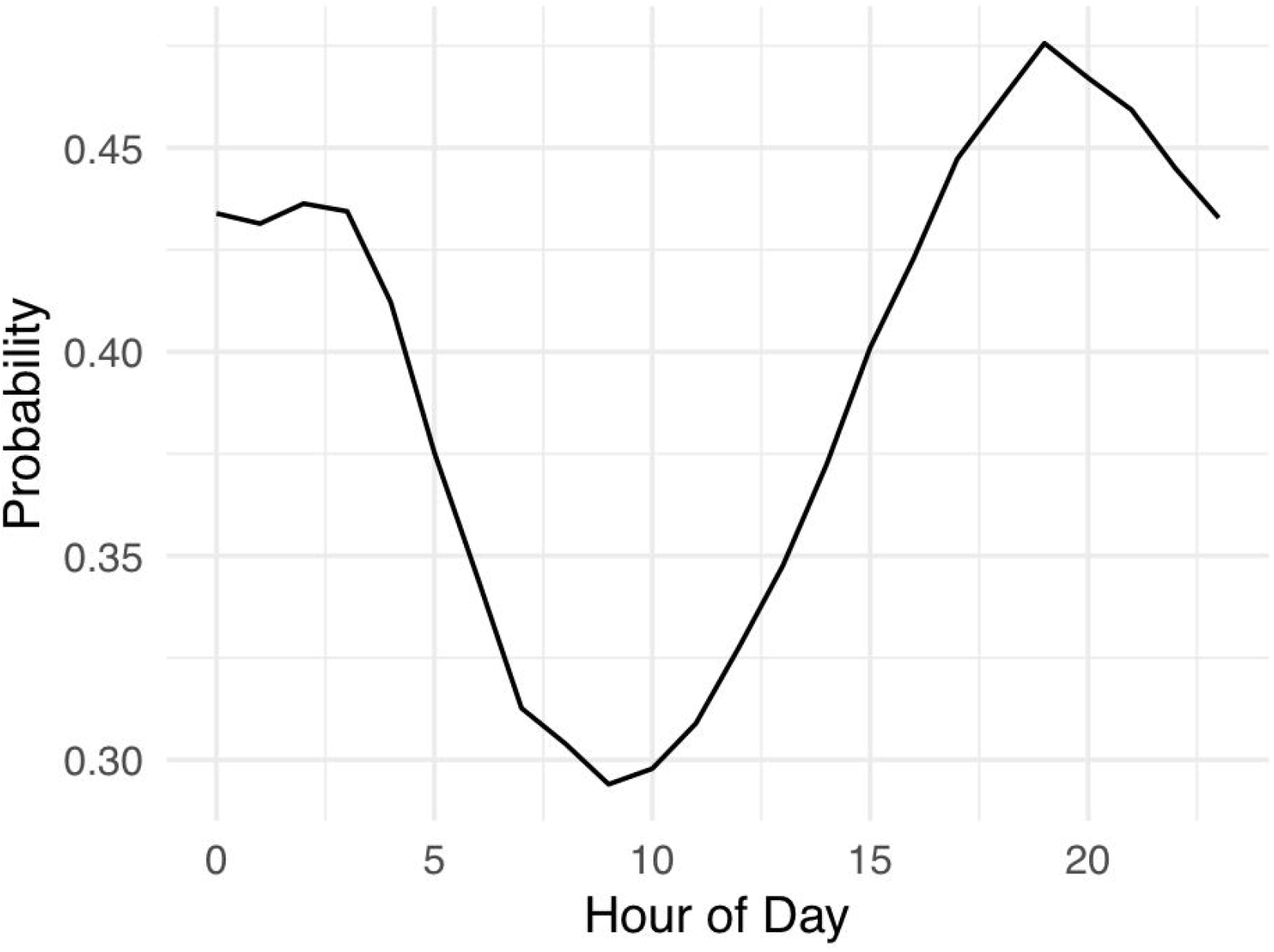
Probability of recording a fever for a representative healthcare worker by time of the day.

### Sensitivity analysis

In order to analyze the sensitivity of our results to the way in which we define an illness episode, we perform a sensitivity analysis by varying the cutpoint to determine the end of an illness episode. To do so, we vary the cutpoint from 24 hours, and repeat both our mean temperature analysis and fever probability analysis with cut-points of 12, 36, 48, and 72 hours. Model predictions for each of these cutpoints can be seen in **Appendix Figures 4-7**.

While there are slight variations in the model predictions when varying the cut-point used to identify an illness episode, as seen below, there are no significant departures from the patterns observed and described for the 24-hour cut-point model. The general pattern remains consistent across sex and age groups, and the probability of drawing a fever is roughly the same across specifications. Thus, the findings are robust to different specifications of this cut-point.

## Discussion

In this study, we used a large collection of thermometer readings from smart Bluetooth thermometers to demonstrate the variation in temperature readings among subjects during a febrile episode. We confirmed that temperature patterns follow a distinct diurnal pattern, and thus the likelihood of recording a fever (e.g., temperature >100°F), also varies. The likelihood not only varies by time, but also by age and sex. Given the widespread efforts for using fevers to screen populations for the presence of disease during public health emergencies (e.g., COVID-19 and SARS), our results have important public health and clinical implications because most temperatures are recorded only at a single point in time.

The diurnal variation of temperature has been known for hundreds of years. However, most of these fever studies have been focused on hospitalized patients. While some have been focused on ambulatory populations (e.g., school children), our population is the largest non-hospitalized population to date. Our population covers a broad range of ages, geographic locations and illnesses. Despite the fact that the diurnal pattern of fevers exists, this is not widely appreciated in clinical, or especially public health practice. Furthermore, while prior work describes the diurnal patterns, our work provides population-based estimates of the probability, given a febrile episode, that an individual of a given age and sex may present with fever.

Our results have three practical public health implications. First, temperature screenings have been used in workplace environments to help identify contagious individuals, especially during the 2020 COVID-19 pandemic but they were also used in prior public health emergencies. Our modeling results highlight the limitations of detecting a fever during febrile episodes for different populations, especially if screening is only performed in the morning. For example, when screening individuals over the age of 65, only 30% registered a fever in the early morning during a febrile episode. Accordingly, a fixed cutoff of, e.g., 100.4°F, and a single point in time (in the morning at the start of a work shift) may not be sufficient for detecting a fever. Screening at multiple times per day or outside of work hours might help to compensate for settings in which there is a lower probability of recording a fever. Indeed, recommendations for monitoring of temperature, in healthcare workers and patients, at multiple points in time already exist.^14,15^ Our results highlight the importance of these practices.

A second implication of our results relates to recommendations for children returning to school and adults returning to work after being ill. Several public health departments recommend a 24-hour fever-free period before returning to school or work. We demonstrate the importance of checking fevers in the evening to ensure that subjects are truly afebrile.

Stressing the importance of afternoon and evening thermometer readings are also important in clinical settings. For example, following orthopedic surgeries involving hardware placement, patients are routinely instructed to take their temperature following discharge from the hospital. Evening readings may substantially increase the chances of documenting a fever and thus diagnosing a surgical site infection sooner. Finally, the recommendation for multiple readings, including evening readings, could be extended to all populations where checking temperature is an important part of helping to determine if a patient has an infection, necessitating further work-up. Such patients should be instructed to take their temperatures again, in the afternoon or evening, as the probability of detecting a fever will be higher.

This study has a number of limitations. First, the febrile episodes we studied are constructed based on patterns of temperature taking activity. We cannot validate the true illness periods in the fevers we observe and some of the non-febrile episodes may actually reflect an illness. We also cannot identify the cause of any fevers recorded. However, prior work has shown these febrile episodes to be highly correlated with seasonal influenza activity, and our sensitivity analysis found no major differences among the different criteria we used to define febrile episodes. Second, because our data are anonymized, we cannot confirm that the demographic information entered by users is accurate or that the device is being used by the exact user whose profile is selected. We also cannot confirm if multiple user profiles have been created for a single individual being observed. Finally, our results may not be generalizable to the entire population. The user reported demographics are younger and have a greater percentage of female users than the general population. Also, users of Bluetooth thermometers may be more health conscious or more likely to have children.

## Conclusions

Using millions of anonymously collected fever readings, this study demonstrated the wide variability in temperatures recorded during periods of febrile illness. Like a number of prior investigations, we found a consistent diurnal pattern in temperature readings. In general, older individuals recorded lower average temperatures throughout the day and temperature differences were observed between men and women for teenagers, young and middle-aged adults. Our results have practical implications for both clinical and public health practice and in any setting where fever is being used for screening purposes in both healthcare (e.g. clinics and hospitals) and non-healthcare (e.g., airport, workplace, school) settings.

## Data Availability

Data used for this study were provided by Kinsa Inc. Availability and access to these data was granted by Kinsa Inc and may only be obtained by agreement with Kinsa Inc.

https://www.kinsahealth.co/contact-us/

## Notes

### Competing Interest Statement

The authors have declared no competing interest.

### Funding Statement

No external funding was received for this work

### Author Declarations

This research uses anonymously collected and de-identified data and is considered IRB exempt by the University of Iowa.

